# Relationship between odor intensity estimates and COVID-19 population prediction in a Swedish sample

**DOI:** 10.1101/2020.05.07.20094516

**Authors:** Behzad Iravani, Artin Arshamian, Aharon Ravia, Eva Mishor, Kobi Snitz, Sagit Shushan, Yehudah Roth, Ofer Perl, Danielle Honigstein, Reut Weissgross, Shiri Karagach, Gernot Ernst, Masako Okamoto, Zachary Mainen, Erminio Monteleone, Caterina Dinnella, Sara Spinelli, Franklin Mariño-Sánchez, Camille Ferdenzi, Monique Smeets, Kazushige Touhara, Moustafa Bensafi, Thomas Hummel, Noam Sobel, Johan N. Lundström

## Abstract

In response to the COVID-19 pandemic, countries have implemented various strategies to reduce and slow the spread of the disease in the general population. For countries that have implemented restrictions on its population in a step-wise manner, monitoring of COVID-19 prevalence is of importance to guide decision on when to impose new, or when to abolish old, restrictions. We are here determining whether measures of odor intensity in a large sample can serve as one such measure. Online measures of how intense common household odors are perceived and symptoms of COVID-19 were collected from 2440 Swedes. Average odor intensity ratings were then compared to predicted COVID-19 population prevalence over time in the Swedish population and were found to closely track each other (r=-0.83). Moreover, we found that there was a large difference in rated intensity between individuals with and without COVID-19 symptoms and number of symptoms was related to odor intensity ratings. Finally, we found that individuals progressing from reporting no symptoms to subsequently reporting COVID-19 symptoms demonstrated a large drop in olfactory performance. These data suggest that measures of odor intensity, if obtained in a large and representative sample, can be used as an indicator of COVID-19 disease in the general population. Importantly, this simple measure could easily be implemented in countries without wide-spread access to COVID-19 testing or implemented as a fast early response before widespread testing can be facilitated.

## INTRODUCTION

The coronavirus disease 2019 (COVID-19) is caused by severe acute respiratory syndrome coronavirus 2 (SARS-CoV-2) and has, since its first discovery at the end of year 2019, rapidly spread across countries (Zhu et al., 2020). In sharp contrast to the majority of western European countries, the Swedish authorities have opted to not close down the majority of society as a response to the COVID-19 pandemic. Instead, the Swedish authorities have, in a step-wise fashion, limited the curtailing of normal individual rights to basically four major actions: international travel ban, a ban on public gatherings of more than 50 individuals (down from an initial ban of >500 individuals); forced universities and high schools (students aged above age of 16) to switch to online teaching, and recently, a mandate that restaurants and bars offer table service only (Swedish Public Health Authority, 2020). In addition, the Swedish Public Health Authority has recommended that individuals work from home, if possible, and has promoted the practice of physical distancing; recommendations that are largely respected. The Swedish response to the pandemic can be summarized as a tactic of step-wise implementation of measures in response to the predicted prevalence of COVID-19. This strategy as a response to a pandemic is, however, dependent on the non-trivial problem of identifying a reliable measure of COVID-19 positive individuals in the population, data of vital importance for the timing of government action to slow the spread of the disease.

Recent reports have demonstrated that inquiries on search engines for salient COVID-19 symptoms can serve as a potential indicator of COVID-19 prevalence in the population (Walker et al., 2020). However, the clinical symptoms of COVID-19 are diffuse (World Health Organization, 2020) which limits the precision of online searches. While the list of identified symptoms has evolved, several recent reports suggest that olfactory dysfunction may be a specifically salient and potentially early symptom of COVID-19 (Gane et al., 2020; Menni et al., 2020). If so, olfactory dysfunction might be a key symptom that can potentially be used as an efficient tool to estimate the prevalence of COVID-19 positive cases in a population. The reported prevalence of olfactory dysfunction in relation to COVID-19 has ranged from around 5% (Mao et al., 2020) to a full 98% (Moein et al., 2020). Reports based on retrospective review of medical records have generally reported lower prevalence numbers, whereas the majority of publications using self-reported olfactory problems have indicated prevalence numbers around 60% (ranging from 30% to 88%) (cf. Pellegrino et al., 2020). This large variation between studies can be attributed to both difference in how olfactory dysfunction was assessed and defined, as well as what sample (clinical or general population) was recruited to the various studies. It is not surprising then that studies only including COVID-19 patients demonstrate larger prevalence of olfactory dysfunction compared to studies with an open inclusion, irrespective of established diagnosis or type of symptoms experienced. Nonetheless, there are currently several peer-reviewed and non-peer-reviewed reports that consistently indicate that olfactory dysfunction is a salient symptom of COVID-19 (cf. Pellegrino et al., 2020). Importantly, the Swedish Public Health Authority recently identified olfactory dysfunction as the most prevalent symptom in the Swedish COVID-19 population estimate. Taken together this indicates that olfactory dysfunction is a good marker for COVID-19 occurrence.

The relationship between self-assessed and psychometrically assessed olfactory function is, however, low (Landis et al., 2003). While most people can notice sudden changes in olfactory function, awareness of an olfactory loss is still far lower than a perceptual loss in other sensory modalities, such as audition and vision. It is therefore necessary to reliably estimate olfactory loss by probing olfactory functions with actual odors. The majority of clinical tests for odor dysfunction use an easily administrated and time-efficient method of assessing an individual’s ability to identify odor (Kobal et al., 1996). However, to create odor identification tests for home use is a problematic undertaking given that the tested individual should not know which odors are included in the test, that test-retest learning is a considerable confound, and that because free odor identification is difficult even for normosmic individuals, written or verbal cues in combination with lures are needed. Together, these problems render odor identification sub-optimal as a home test. A more straightforward test might therefore be to assess odor intensities of common household odors, thereby avoiding the limitations imposed by verbalization. Indeed, using assessment of odor intensities as a measure of olfactory functions has previously been used (Stamps et al., 2013) and demonstrated to be related to odor detection thresholds of the odor in question (Kern et al., 2015).

Here we will assess whether odor intensity ratings can be used as a measure of COVID-19 spread in the population. We do this by using data from a Swedish population to determine whether a home-based odor intensity rating can predict population prevalence of COVID-19 in Sweden. This data comes from a multicenter project initiated at the Weizmann Institute of Science with the overall aim of determining the involvement of the olfactory system in the COVID-19 disease. Via an online rating tool (www.smelltracker.org), participants rate household odors for their perceived intensity and pleasantness. Specifically, we determine whether odor intensity ratings negatively follow predicted population prevalence of COVID-19 and whether odor intensity ratings are modulated by COVID-19 symptoms. We hypothesized that, in a larger sample, odor intensity ratings over time would follow the prevalence of COVID-19 in the general population and therefore be of potential use as a predictor.

## METHODS

### Participants

A total of 2930 unique individuals entered the data collection website *smelltracker.org*, identified themselves as Swedish, and provided information about their sex and age. In our analyses, we removed: 33 individuals that indicated an age below 18, 374 individuals who did not provide any odor ratings, and 83 individuals that rated all odor intensities above 95 (on a 0-100 scale, see below for scale information) on suspicion of not following/understanding the task. This meant that our final sample consisted of a total of 2440 individuals (mean age 47.4 years, ±14.11 SD, range 18-99). In the final sample, a total of 1680 individuals identified themselves as a woman and 760 as a man. Data collection and analyses were approved by both the Israeli Edith Wolfson Medical Centre Helsinki Committee and the Swedish Institutional Review Board (Etikprövningsnämnden, 2020-01577).

Participants did not receive any form of monetary compensation for their participation and consent was waived. All aspects of the study complied with the Declaration of Helsinki for Medical Research involving Human Subjects.

### Recruitment strategy and participant regional domiciliation

Recruitment for the Swedish population was mainly done by appearances in various local news media in the Stockholm regions and outreach initiatives by the Karolinska Institute in Stockholm. Due to the high level of privacy security, no data that could localize the participant other than to country of residency was obtained; we therefore do not know with certainty in what region of Sweden participants were localized at the time of testing. However, judging from COVID-19 related hospitalization rates in the Swedish regions at the time of sampling, the spread of COVID-19 has mostly been centered around the general Stockholm area. The regional specificity in both COVID-19 spread and recruiting, in combination with the demonstrated association between our obtained data and COVID-19 prediction in the Stockholm area (see Results), we believe that the vast majority of participants were from the greater Stockholm area.

### Procedure

Participants visited the Swedish version of the multilingual website smelltracker.org and provided details regarding age, declared gender (Woman/Man/Other), and whether they have been tested for COVID-19 (No, Yes-Pending, Yes-Positive, Yes-Negative). They subsequently answered what symptoms of COVID-19, if any, they currently experienced. Available symptoms were: Fever, Cough, Shortness of breath or difficulty breathing, Tiredness, Aches, Runny nose, Sore throat, Loss of the sense of smell, Loss of taste, No symptoms. Unique login was created for each individual to facilitate repeated testing.

Next, participants picked 5 odors to rate, each from a separate category with a fixed list of common household odors. The first two odor categories were selected to contain odors with little to no trigeminal sensation (unisensory odors) whereas the last 3 categories where odors with mixed sensations of odor and trigeminal in various degree, so called bimodal odors (Table 1). Participants were instructed to preferentially pick from the top of each list but, if necessary, choose any item that would be available to them going down the list in order. At repeated testing, the same 5 odors, freshly prepared, were to be used. Participants then proceeded to smell each odor and, on a separate page for each odor, rated their perceived intensity and pleasantness on separate visual analogue scales ranging from very weak/very unpleasant to very strong/very pleasant, respectively. These scales were coded in the system as ranging from 0 (min) to 100 (max). Participants could smell the odor as often as they liked, and there was no time pressure applied. We are here only focusing on odor intensity ratings.

**Table 1.** Odor categories with the alternatives available for participants to choose from.

| Category 1 | Category 2 | Category 3 | Category 4 | Category 5 |
| --- | --- | --- | --- | --- |
| Vanilla extract | Peanut butter | Mustard (Dijon) | Garlic (chopped) | Toothpaste |
| Nutella | Coconut oil | Vinegar (white) | Camembert cheese | Hand soap |
| Honey | Olive oil | Horseradish (jar) | Canned tuna | Laundry detergent |
| Strawberry jam | Basil | Wasabi | Blue cheese | Shampoo |
| Apricot jam | Oregano | Onion (chopped) | Canned sardines | Hand cream |
| Apple juice (not fresh) | Parsley | Vinegar (apple) | Mushrooms | Body lotion |
| Orange juice (not fresh) | Cilantro | Black pepper (ground) | Boiled egg | Perfume |
| Lemonade (not fresh) | Dill | Menthol gum | Pickled herring | Hand sanitizer |
| Peach nectar (not fresh) | Cardamom | Mint (fresh) | Cumin | Sunscreen |
| Pear nectar (not fresh) | Thyme | Mint (gum) | Soy sauce | Baby oil |
| Grapefruit juice (not fresh) | Nutmeg | Mint (tea) | Sauerkraut (jar) |  |
| Pineapple juice (not fresh) | Caraway | Sesame oil | Coffee (ground) |  |
| Banana nectar (not fresh) | Bay leaves | Vodka | Coffee (instant) |  |
| Cinnamon | Ketchup | Clove | Tea (black) |  |
| Maple syrup | Peanut butter | Vinegar (balsamic) | Tea (earl grey) |  |

*Note. Categories 1-2 contain items with odors that are low in trigeminal irritation whereas categories 3-5 contain odors with a higher trigeminal irritation factor.*

### Population prevalence and palynological data

We obtained data on the predicted prevalence of COVID-19 in the sample population from the Public Health Agency of Sweden (Folkhälsomyndigheten). From March to April, Folkhälsomyndigheten randomly sampled 738 individuals in the Stockholm region and based on this data, together with available data from the healthcare system and the contagion factor of the SARS-CoV-2 virus, modelled the prevalence of COVID-19 in the Stockholm population over time. The model is a fitted compartmental SEIR model that assume unreported cases as 98.7% of infected and their infectivity as 55% compared to reported cases. Model details, raw data, scripts, and figures for the updated model (version 2), that were used as a predictor in this manuscript, can be obtained from an open data deposit: https://github.com/FohmAnalys/SEIR-model-Stockholm

We also obtained airborne pollen data for the Stockholm area from the Palynological laboratory at the Swedish Museum of Natural History. Because we do not have data on the prevalence of potential allergic rhinitis (pollen allergy) or what specific allergy participants had, we summed up the values of all allergenic pollen for each day of measuring. During the time period of our sampling, there were airborne pollen detected from three species; Alder (Alnus), Birch (Betula), and Hazel (Corylus).

### Data reduction and statistical analyses

To match the results of the COVID-19 prediction model and to increase the reliability of the collected intensity ratings, values were averaged in 3-day intervals. Single cases of missing values were replaced with median of that specific odor category. Odor ratings were then averaged across odors. For analyses of global odor intensity, all categories were averaged; for ratings of unisensory odors, categories 1-2 were averaged, and for ratings of bimodal odors, categories 3, 4, and 5 were averaged

Associations between odor intensity ratings and COVID-19 prediction model, as well as odor intensity and COVID-19 symptoms, were assessed by Spearman rank correlations. To identify olfactory dysfunction threshold, a cut-off of the 10th percentile of odor intensity within the subpopulation that reported no symptom were used as an indication of olfactory symptoms, as is often used for clinical olfactory tests (Hummel et al., 2007). This cut-off value was subsequently used to define percentage of olfactory dysfunction in other subpopulations, i.e. (subjectively reported) Symptom, COVID-19 + and COVID-19 -, the last two categories based on lab testing. To maximize likelihood of identifying individuals with COVID-19 tests, in these analyses, we included all sessions from individuals with repeated testing.

To assess ability of our measure to identify shift in potential diagnose, we identified individuals who had provided data on more than one occasion and from them, assessed who had progressed from indicating no symptoms to indicating symptoms in a subsequent session. Their ratings in the session just before the session indicating symptoms were used as their non-symptomatic score and their ratings in the first session indicating symptoms were used as their symptomatic scores. A paired two tailed Student’s t-tests were used to assess odor intensity rating pre- and post-developing COVID-19 symptoms. Finally, for the test-retest reliability, we used Pearson correlation between odor intensity ratings in the first session and second session in individuals who provided data in more than 1 session and who reported no symptoms in both. All other correlation tests were done with Spearman correlation to avoid influence of skewed data. All statistical analyses were carried out within the MATLAB (version 2019b) environment with Statistical and Machine Learning toolbox.

## RESULTS

### Relationship between COVID-19 prevalence and odor intensity perception

We assessed development of olfactory abilities over time by plotting mean intensity perception per day against the predicted prevalence of COVID-19 in the Stockholm population. Over the testing time, mean odor intensity ratings decline whereas the predicted COVID-19 societal levels go up (Figure 1A). there was a marked downward shift in ratings of odor intensity levels occurring in this sample between the 4th to 9th of April. To assess whether this function was different for odors with low and high trigeminal irritants (unisensory and bimodal odors), we assessed them separately. We found that both the unimodal odor category (Figure 1B) and the bimodal odor category (Figure 1C) had similar psychometric functions. We then assessed the statistical relationship between intensity estimates over time and population COVID-19 predictions using separate Spearman rank correlation. There was a significant negative relationship between the COVID-19 prediction model and odor intensity ratings over time, *ρ* = - 0.83, *p* < .001 (Supplementary Figure S1). Similarly, there were significant relationships between the COVID-19 prediction model and both the unimodal odor category, *ρ* = −.79, *p* < .003, as well as the bimodal odor category, *ρ* = −0.83, *p* < .001. Indeed, in a separate model, the Swedish Public Health Authorities predicated that peak prevalence of individuals that were contagious with the SARS-CoV-2 virus in the Stockholm area occurred on the 8th of April (Swedish Public Health Authorities, 2020). In other words, odor intensity ratings of common household odors track estimated prevalence of COVID-19 in the population and the shift in intensity aligned with predicted peak SARS-CoV-2 contagion. This seems independent of whether the odors have more or less trigeminal irritants which is consistent with the high correlation between ratings of the unisensory and bimodal odors, *ρ* = 0.92, *p* < .001.

**Figure 1.**
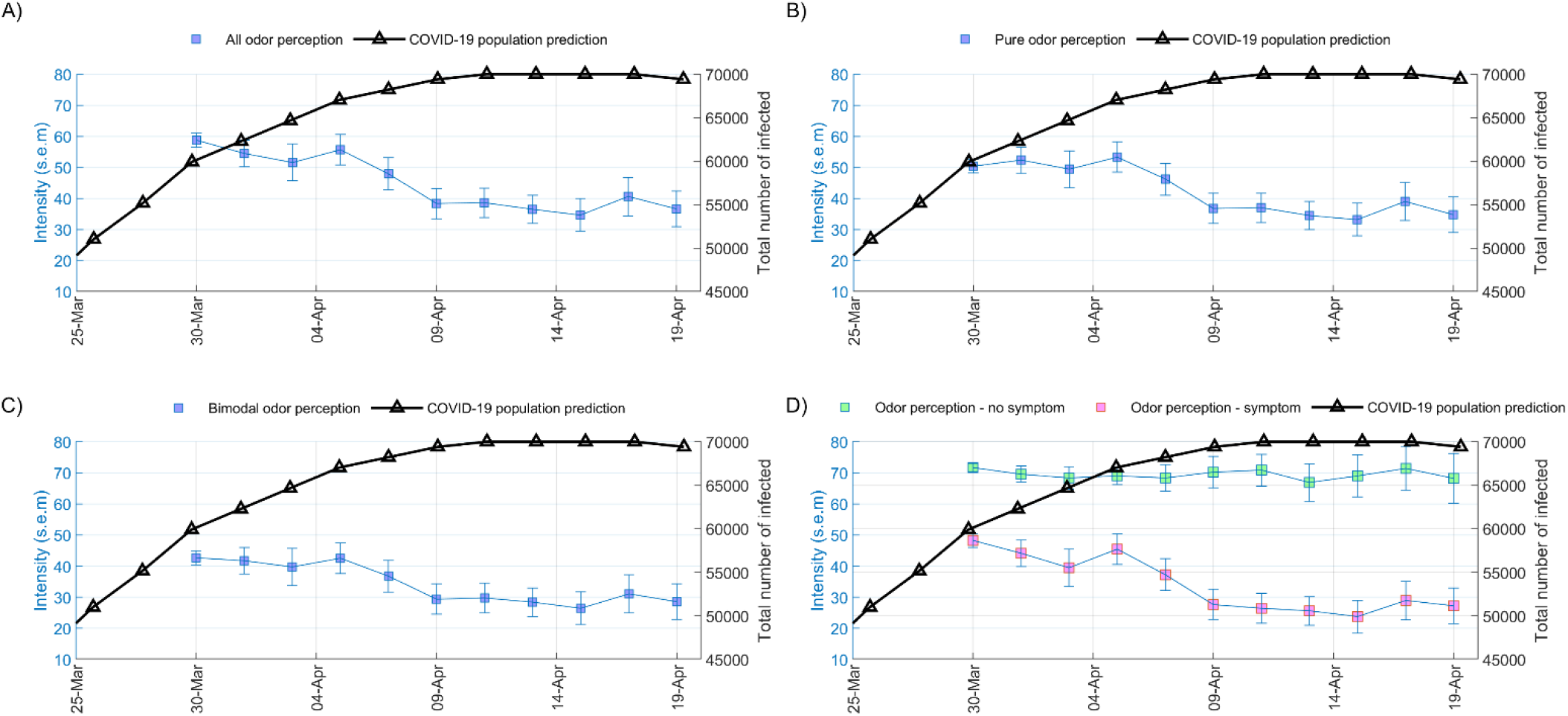
Odor intensity perception relate to COVID-19 prevalence. **A**) Mean intensity ratings of the 5 odor categories (blue line and axis) in relation to population prediction (black line and axis) of COVID-19 prevalence in the Stockholm region. **B**) Mean intensity ratings of unimodal odors (odor categories 1-2; blue line and axis) in relation to population prediction of COVID-19 prevalence in the Stockholm region. **C**) Mean intensity ratings of bimodal odors (odor categories 3-5; blue line and axis) in relation to population prediction of COVID-19 prevalence in the Stockholm region. **D**) Mean intensity ratings of odors (category 1-5), separated into individuals without (green squares, blue axis) and with (purple squares, blue axis) reported COVID-19 symptoms, in relation to population prediction (black line and axis) of COVID-19 prevalence in the Stockholm region. Error bars in all panels indicate standard error of the mean (s.e.m).

In this sample, as in the general population, there are healthy individuals who might have done the odor testing as a potential screening tool for COVID-19. We therefore wanted to assess the potential influence of COVID-19 symptoms on odor intensity ratings. To this end, we first divided the sample into individuals who reported no COVID-19 symptoms and individuals who reported at least one of the listed symptoms. There was a clear separation in odor intensity perception between the two groups across time (Figure 1D). Moreover, the more symptoms the individual reported, the weaker the odors were perceived as, as demonstrated by a Spearman correlation between number of reported COVID-19 symptoms and odor intensity ratings, *ρ* =-.29, *p* < .001 (Fig 2).

**Figure 2.**
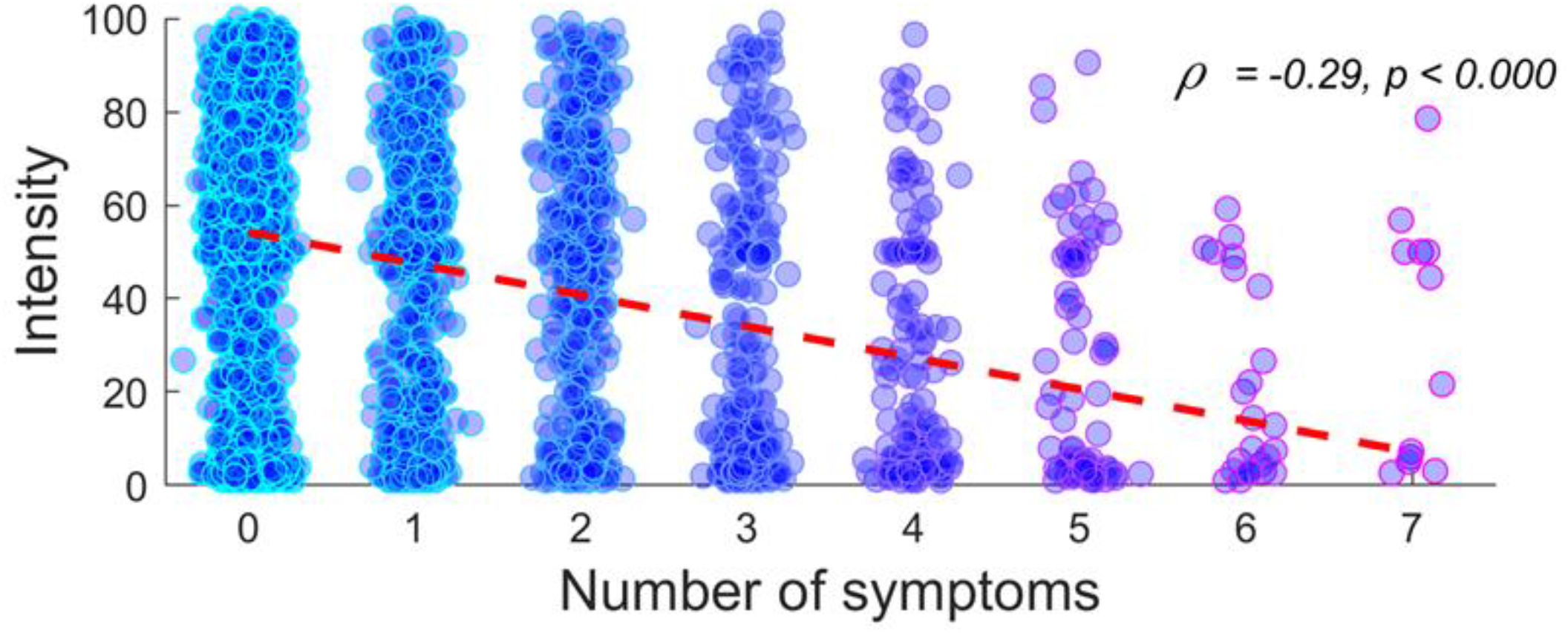
Odor intensity perception relates to COVID-19 symptoms. Individual mean rated intensity of odors in relation to number of reported COVID-19 symptoms, excluding loss of smell/taste. Dots represent individuals and red dotted line indicates the regression line. Blue color indicate number of overlapping individuals.

A potential confounding variable in this Swedish sample is the co-occurrence of the COVID-19 pandemic with the onset of the Swedish pollen season. In other words, a potential shift in rated intensity over time could be mediated by an increase in allergic rhinitis due to a rise in levels of airborne pollen. We therefore plotted the change in odor intensity ratings over time compared to daily summarized levels of measured airborne pollen from all the known allergenic species detected at the time in the Stockholm area. There was a clear increase in summated pollen levels towards the end of our sampling period (Supplementary Figure S2). However, this increase in summated pollen levels occurred at a later time-point than the marked downward shift in ratings of odor intensity levels occurring in this sample between the 4th to 9th of April.

### Higher of prevalence of olfactory dysfunction among individuals with COVID-19 symptoms

Having established that there is a clear difference in odor intensity perception between individuals with and those without COVID-19 symptoms, we wanted to know what the prevalence of olfactory dysfunction were in the overall sample and the different subsamples. We defined olfactory dysfunction as mean odor intensity ratings falling below the 10th percentile in the group reporting no COVID-19 symptoms; a definition that is aligned with other attempts of assessing prevalence of olfactory dysfunction (Hummel et al., 2007). This is of course a conservative measure that will bias our estimates towards a potential lower prevalence given that there might be asymptomatic COVID-19 patients or individuals with olfactory dysfunction prior to the COVID-19 pandemic. Applying this cut-off to the group that displayed symptoms of COVID-19, 66% of these individuals had intensity estimates falling in the olfactory dysfunction category (Figure 3A). However, having symptoms of COVID-19 does not constitute a diagnosis meaning that odor dysfunction in the sample cannot be directly linked to confirmed COVID-19 diagnose. The national strategy of the Swedish national healthcare system has been to prioritize testing of individuals admitted to a hospital with clear signs of a SARS-CoV-2 infection. This means that few individuals in our sample had been tested for COVID-19 and even fewer had a positive test. However, 16 individuals indicated that they had been confirmed as COVID-19 and among these individuals, 81% was classified in the olfactory dysfunction group (Figure 3A). Among the individuals that had in the past been confirmed as COVID-19 negative, 32% were classified in the olfactory dysfunction group.

**Figure 3.**
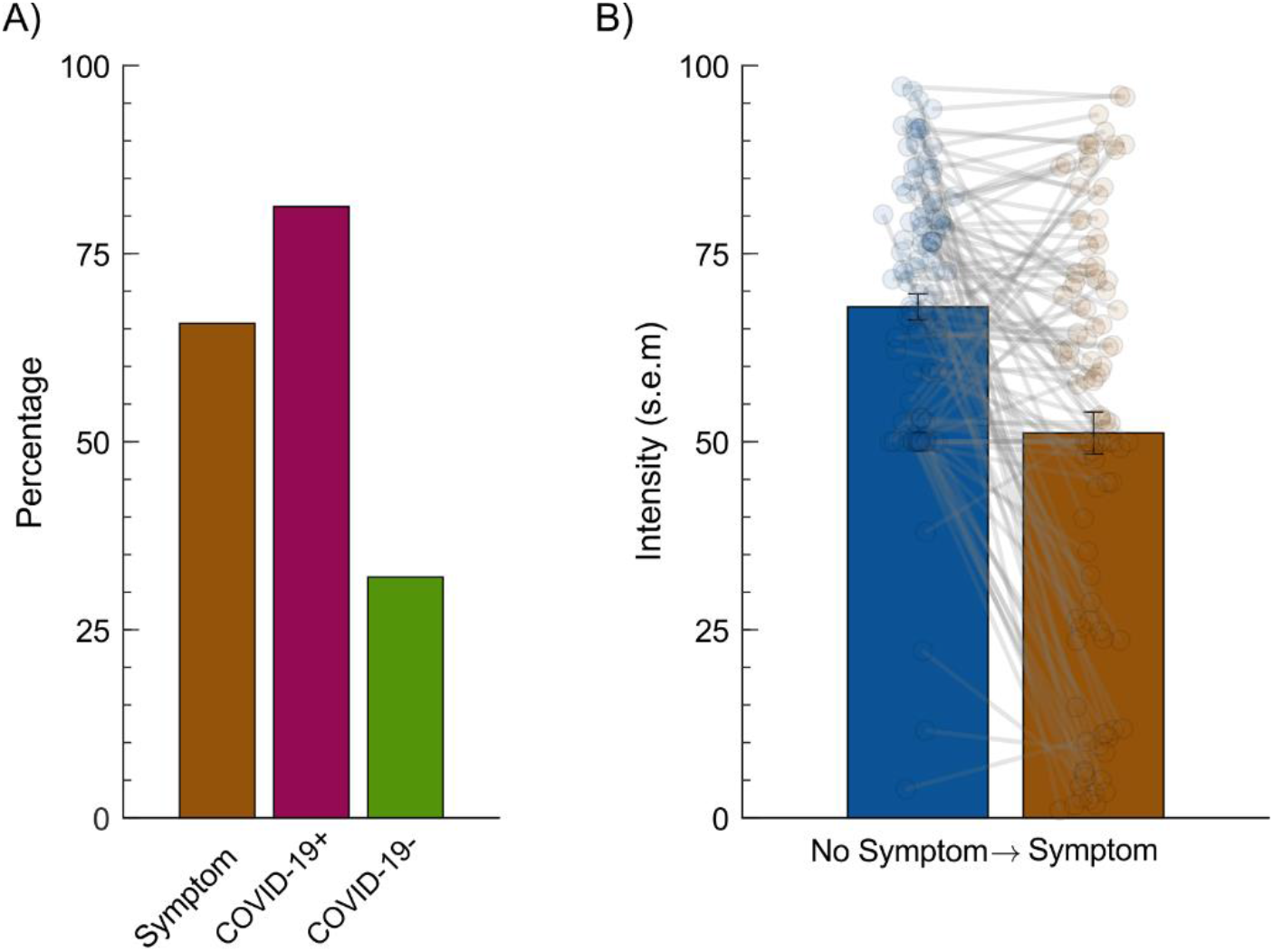
Olfactory dysfunction in relationship to COVID-19 symptoms. **A**) Percentage of olfactory dysfunction within subsample that indicated either COVID-19 symptoms (sessions, n=2469) or had undergone COVID-19 testing (Covid-19 + = positive [n=16], Covid-19 - = negative [n=25]). **B**) Shift in intensity ratings between sessions for individuals that progressed from indicated “No Symptoms” to indicating “Symptoms”. Dots indicate individual values (n=107) and lines connects the values for the same individual. Error bars indicate standard error of the mean (s.e.m).

A subsample visited the website on multiple occasion and provided intensity estimates on repeated occasions. This provided us the opportunity to observe when individuals progressed from indicating “No Symptoms” to indicating one or more symptoms in subsequent testing (Figure 3B). A total of 107 individuals made this transition at some point during testing. We then compared their mean intensity rating of the unisensory odors during the last session they indicated No Symptoms with that of the first session they started to indicate symptoms. On the group level, there was a significant difference between the two sessions, *t*(103) = 6.15 *p* < .001, with an average of 20 points (29%) reduction in odor intensity on the 100 point visual analogue rating scale.

### Test-retest reliability

Finally, we determined the test-retest reliability of the odor intensity measure by assessing the relationship between odor ratings in their first session and their second session within individuals that performed more than 1 testing session and who reported no symptoms in either one of the sessions (n = 130). A Spearman correlation test indicated a decent test-retest coefficient of .66, *ρ* (128) = .66, *p* < .0001.

## DISCUSSION

We can here demonstrate that ratings of odor intensity from a larger group within an area of COVID-19 outbreak closely follows the predicted prevalence of COVID-19 over time. We can also demonstrate that not only is odor dysfunction associated with symptoms of COVID-19, but that when an individual progress from indicating no COVID-19 symptoms to listing COVID-19 symptoms, there is a significant drop in odor intensity as soon as the next testing session. These results suggest that simple perceptual ratings of odors can serve as a future tool to predict levels of COVID-19 infection within a population.

Home testing of odor functions during a pandemic need to be easy both to implement and to explain to the individual. Intensity ratings of widely available household products via online assessment is a simple and cheap way both for the individual to obtain indication of odor dysfunction, which might indicate COVID-19 onset, and for a potential governmental or health organization to monitor spread of the disease. Participants in this study had to choose between set classes of odors, but this may not be necessary if the main aim is to determine COVID-19 spread based on intensity estimates. Intensity ratings for the odor categories containing odor sources with low trigeminal irritants and the categories containing odor sources with a bimodal sensation both closely followed the predicted COVID-19 prevalence (correlation coefficient .79 and .83, respectively). Few household odors beyond vanillin are truly activating only the olfactory system without any trigeminal activation and attempting to only use low-trigeminal odors might lower the ease of use; therefore, a more feasible setup might be to allow free selection and instead block certain odor sources with a known high trigeminal activation. It is at the present time unclear if the SARS-CoV-2 virus affects the olfactory system, alone, or whether the trigeminal system is also influenced. The fact that bimodal odors, as well, closely track the COVID-19 spread in the population lends support to the notion that the trigeminal system may also be affected, either by the SARS-CoV-2 virus or by the reduced olfactory ability per se (Frasnelli et al., 2007). However, although the odors listed in the bimodal odor categories are bimodal in their percept, most are not strong trigeminal irritants. Whether similar results would be obtained using stronger irritants, such as acetone etc., remains to be determined.

Of the 16 individuals with confirmed COVID-19, 81% had intensity ratings low enough to fall in the olfactory dysfunction category. Even though this result is higher than what have been reported in studies based on self-reports, this high number is aligned with a recent study that tested actuall olfactory function in confirmed COVID-19 positive patients (Moein et al., 2020). At the time of the study, the Swedish health authorities prioritized testing of COVID-19 in individuals when admitted to a hospital with signs of COVID-19 and key healthcare staff. This has led to a low number of individuals with confirmed COVID-19 status which is a major weakness of the study. We can therefore not conclusively state that all individuals with symptoms of COVID-19 indeed were COVID-19 positive. We did find, however, that number of listed symptoms was negatively associated with odor intensity and there was, on average, a large drop in odor intensity perception when an individual transgressed from reporting no symptoms to reporting symptoms.

Still, it is important to note that a shift in odor intensity perception is not by itself a diagnosis of COVID-19 and future studies addressing links between COVID-19 and olfactory perception should, if possible, test individuals with established COVID-19 status to obtain exact measure of its specificity. That said, a major benefit of the study is the demonstration that ratings of odor intensities do track COVID-19 population prevalence which could potentially be of use in countries where in-field testing is not available or at initial stages of potential future outbreak of corona virus that might have similar effects on the olfactory system and where the development and buildup testing capacity has yet to happen.

The marked drop in olfactory functions between sessions when participants started to report COVID-19 symptoms suggests that odor measures might serve as a clear indicator of COVID-19 at an individual level beyond the population level demonstrated here. However, the test-retest reliability of odor intensity measure, here estimated to .66, indicate that odor intensity function might not work as a reliable measure on the individual level. Indeed, other studies assessing test-retest of odor intensities have reached a near identical value (Kern et al., 2015). Hence, to achieve a high reliability at the individual level, a more general and advanced odor test than mere intensity ratings is probably needed to have the specificity needed. At this time, it is unlikely that mere intensity estimates could serve as anything other than an indication of a potential COVID-19 diagnosis which should either trigger serum testing of COVID-19 or for healthcare workers to treat a patient as a potential COVID-19 suspect. Nonetheless, there is great potential for the development of odor tests of COVID-19 that might reach a specificity near serum tests.

In the general debate whether olfactory dysfunction is a sign of COVID-19, a valid counter argument has been raised; that loss of olfactory function is not uncommon for upper respiratory infections and the significantly larger prevalences of olfactory dysfunction reported for the SARS-CoV-2 virus might be explained by the increased attention to symptoms and an increase in response prevalence. Upper respiratory infections do commonly lead to a reduction of olfactory function or anosmia (total loss of olfactory functions) (Seiden, 2004) and virus infection is one of the leading causes of anosmia in patients with nonconductive olfactory disorders (Quint et al., 2001). However, an interesting aspect of COVID-19 related olfactory dysfunction seems to be the frequent report of an isolated sudden onset in absence of other nasal problems with patients reporting a normal nasal patency (Gane et al., 2020). The exact mechanism of this onset of olfactory dysfunction without other common symptoms associated with upper respiratory infections has yet to be determined but recent data suggest a few hypotheses. Nasal epithelial cells richly express the SARS-CoV-2 entry factor (Sungnak et al., 2020), the ACE2 receptors, which has been linked to both the virus replication rate and disease severity (Hoffmann et al., 2020; Zhou et al., 2020){ref}. Based on this, it can be speculated that the SARS-CoV-2 virus damage the olfactory epithelium to such extent that olfactory ability is degraded. Many viruses, including corona viruses, can also propagate via the olfactory nerve and thereby infect and damage the olfactory bulb (Schwob et al., 2001; Wheeler et al., 2017). Corona virus RNA has been found in olfactory areas of the brain (Li et al., 2020), and recent data suggests that some COVID-19 patients display neurological symptoms (Mao et al., 2020). Moreover, it is possible that nasal blockage occur but so high up in the nasal cavity that the blockage only affects access to the olfactory epithelium without impeding nasal patency (Trotier et al., 2007). Future studies are needed to isolate the exact mechanisms.

In conclusion, we can here demonstrate that measures of odor intensity closely track estimated COVID-19 levels on a population level. This simple measure, if implemented in a large sample within the area of outbreak, could serve as an easy and cheap measure of COVID-19 spread in society. This measure would provide special value to underdeveloped countries where COVID-19 tests might not be widely available or to be implemented in an early phase of a COVID-19 epidemic before wide-spread testing has been implemented.

## Data Availability

Model details, raw data, scripts, and figures for the updated model (version 2), that were used as a predictor in this manuscript, can be obtained from an open data deposit: https://github.com/FohmAnalys/SEIR-model-Stockholm

## ACKNOWLEDGEMENTS

This project was supported by an ERC AdG grant (SocioSmell, 670798), awarded to Noam Sobel, and the writing and analyses by a grant from the Knut and Alice Wallenberg Foundation (KAW 2018.0152), awarded to Johan N. Lundström.

